## Supplementary material for "A Core Outcome Set to evaluate the impact of prognostication in people living with advanced cancer: an international consensus study": S1 File

### Additional demographic data

| Patient characteristics | Round 1 (*n* = 10) | Round 2 (*n* = 4) | Consensus meeting (*n* = 2) |
|---|---|---|---|
| **Age category (years), n (%)*** | | | |
| 26 - 35 | 1 (10) | 1 (25) | 1 (50) |
| 36 - 45 | 1 (10) | - | - |
| 56 - 65 | 6 (60) | 1 (25) | 1 (50) |
| 66 - 75 | 2 (20) | 2 (50) | - |
| **Gender, n (%)** | | | |
| Male | 4 (40) | 1 (25) | 1 (50) |
| Female | 5 (50) | 3 (75) | 1 (50) |
| Prefer to self-describe | 1 (10) | - | - |
| **Ethnicity, n (%)** | | | |
| White (British) | 5 (50) | 3 (75) | 2 (100) |
| White (Southern Balkan) | 1 (10) | - | - |
| White (Swiss) | 1 (10) | - | - |
| White (Not specified) | 1 (10) | - | - |
| Other (Not specified) | 1 (10) | - | - |
| Prefer not to say | 1 (10) | 1 (25) | - |
| **Country of residence, n (%)** | | | |
| England | 6 (60) | 4 (100) | 2 (100) |
| Scotland | 1 (10) | - | - |
| Switzerland | 2 (20) | - | - |
| Wales | 1 (10) | - | - |
| **Patient type, n (%)** | | | |
| Inpatient | 1 (10) | - | - |
| Outpatient | 9 (90) | 4 (100) | 2 (100) |
| **Site of primary cancer, n (%)** | | | |
| Abdominal | 1 (10) | 1 (10) | - |
| Liver | 1 (10) | 1 (10) | 1 (50) |
| Lung | 2 (20) | - | - |
| Lymphatic/haematological | 1 (10) | - | - |
| Multiple myeloma | 1 (10) | 1 (10) | - |
| Prostate | 1 (10) | - | - |
| Soft tissue sarcoma | 1 (10) | 1 (10) | 1 (50) |
| Stomach | 2 (20) | - | - |
| **Highest educational qualification, n (%)** | | | |
| Secondary education | 1 (10) | - | - |
| Higher education below degree | 1 (10) | - | - |
| Degree | 3 (30) | 2 (50) | 1 (50) |
| Postgraduate | 4 (40) | 2 (50) | 1 (50) |
| No qualifications | 1 (10) | - | - |

| **Informal caregiver characteristics** | **Round 1** (*n* = 11) | **Round 2** (*n* = 5) | **Consensus meeting** (*n* = 2) |
|---|---|---|---|
| **Age category (years), n (%)** | | | |
| 18 - 25 | 1 (9) | - | - |
| 36 - 45 | 2 (18) | 1 (20) | - |
| 46 - 55 | 3 (27) | 2 (40) | 2 (100) |
| 56 - 65 | 2 (18) | - | - |
| 66 - 75 | 2 (18) | 2 (40) | - |
| 76 - 85 | 1 (9) | - | - |
| **Gender, n (%)** | | | |
| Male | 2 (18) | 1 (20) | - |
| Female | 8 (73) | 3 (60) | 2 (100) |
| Prefer not to say | 1 (9) | 1 (20) | - |
| **Ethnicity, n (%)** | | | |
| Black (African) | 1 (9) | - | - |
| Mixed/multiple (Anglo-Burmese) | 1 (9) | - | - |
| Mixed/multiple (Asian and White) | 1 (9) | 1 (20) | 1 (50) |
| Mixed/multiple (Not specified) | 1 (9) | - | - |
| White (British) | 5 (45) | 3 (60) | 1 (50) |
| White (Swedish) | 1 (9) | - | - |
| Prefer not to say | 1 (9) | 1 (20) | - |
| **Country of residence, n (%)** | | | |
| England | 10 (91) | 5 (100) | 2 (100) |
| Sweden | 1 (9) | - | - |
| **Current caring situation, n (%)** | | | |
| Bereaved caregiver | 5 (45) | 2 (40) | 1 (50) |
| Current caregiver | 6 (55) | 3 (60) | 1 (50) |
| **Relationship to patient, n (%)** | | | |
| Child | 1 (9) | - | - |
| Friend | 3 (27) | 2 (40) | 1 (50) |
| Husband/Wife/Partner | 3 (27) | 1 (20) | - |
| Parent | 2 (18) | 2 (40) | 1 (50) |
| Sister-in-law | 1 (9) | - | - |
| Not specified | 1 (9) | - | - |
| **Site of patient's primary cancer, n (%)** | | | |
| Breast | 1 (9) | 1 (20) | - |
| Eye/brain/central nervous system | 1 (9) | - | - |
| Lung | 4 (36) | 2 (40) | 2 (100) |
| Lymphatic/haematological | 1 (9) | 1 (20) | - |
| Metastatic melanoma | 1 (9) | - | - |
| Pancreas | 1 (9) | - | - |
| Prefer not to say | 1 (9) | - | - |
| Unknown | 1 (9) | 1 (20) | - |
| **Highest educational qualification, n (%)** | | | |
| Secondary/high school education | 2 (18) | 2 (40) | 1 (50) |
| Degree | 3 (27) | 1 (20) | - |

| | | | |
|---|---|---|---|
| Postgraduate | 5 (45) | 2 (40) | 1 (50) |
| Prefer not to say | 1 (9) | - | - |
| **Clinician characteristics** | **Round 1** (*n* = 18) | **Round 2** (*n* = 15) | **Consensus meeting** (*n* = 5) |
| **Age category (years), n (%)** | | | |
| 26 - 35 | 3 (17) | - | - |
| 36 - 45 | 4 (22) | 4 (27) | 3 (60) |
| 46 - 55 | 8 (44) | 8 (53) | 1 (20) |
| 56 - 65 | 3 (17) | 3 (20) | 1 (20) |
| **Gender, n (%)** | | | |
| Male | 6 (33) | 5 (33) | 2 (40) |
| Female | 12 (67) | 10 (67) | 4 (80) |
| **Ethnicity, n (%)** | | | |
| Arab | 1 (6) | 1 (7) | - |
| Asian (Chinese) | 1 (6) | 1 (7) | - |
| Asian (Hongkonger) | 1 (6) | 1 (7) | 1 (20) |
| Black (African) | 2 (11) | 1 (7) | 1 (20) |
| White (American) | 1 (6) | 1 (7) | - |
| White (British) | 4 (22) | 3 (20) | 1 (20) |
| White (Canadian) | 1 (6) | 1 (7) | - |
| White (Finnish) | 1 (6) | 1 (7) | - |
| White German) | 1 (6) | 1 (7) | - |
| White (Irish) | 1 (6) | 1 (7) | - |
| White (Portuguese) | 1 (6) | - | - |
| White (Not specified) | 3 (17) | 1 (7) | 2 (40) |
| **Country of residence, n (%)** | | | |
| Belgium | 1 (6) | 1 (7) | - |
| Canada | 2 (11) | 2 (13) | - |
| England | 9 (50) | 8 (53) | 3 (60) |
| Kenya | 1 (6) | - | - |
| Portugal | 1 (6) | - | - |
| Singapore | 1 (6) | 1 (7) | 1 (20) |
| Sweden | 1 (6) | 1 (7) | - |
| Switzerland | 1 (6) | 1 (7) | - |
| USA | 1 (6) | 1 (7) | 1 (20) |
| **Professional role, n (%)** | | | |
| Allied Health professional (Not specified) | 1 (6) | - | - |
| Nurse | 4 (22) | 4 (27) | 1 (20) |
| Occupational therapist | 1 (6) | - | - |
| Physician/doctor | 12 (67) | 11 (73) | 4 (80) |
| **Role location, n (%)** | | | |
| Hospital | 11 (61) | 10 (67) | 4 (80) |
| Hospice | 2 (11) | 1 (7) | - |
| Community | 4 (22) | 4 (27) | 1 (20) |
| Not specified | 1 (6) | - | - |

| | | | |
|---|---|---|---|
| **Highest educational qualification, n (%)** | | | |
| Higher education below degree | 1 (6) | 1 (7) | - |
| Degree | 3 (17) | 1 (7) | - |
| Postgraduate | 14 (78) | 13 (87) | 5 (100) |
| **Length of palliative care experience (years), n (%)** | | | |
| < 1 | 1 (6) | 1 (7) | - |
| 1 - 5 | 4 (22) | 1 (7) | - |
| 6 - 10 | 1 (6) | 1 (7) | 1 (20) |
| 11 - 15 | 7 (39) | 7 (47) | 4 (80) |
| 16 - 20 | 2 (11) | 2 (13) | - |
| > 20 | 3 (17) | 3 (20) | - |
| **Length of healthcare experience (years), n (%)** | | | |
| 1 - 5 | 2 (11) | 1 (7) | - |
| 6 - 10 | 1 (6) | - | - |
| 11 - 15 | 4 (22) | 3 (20) | 3 (60) |
| 16 - 20 | 5 (28) | 5 (33) | 2 (40) |
| > 20 | 6 (33) | 6 (40) | - |
| **Academic/researcher characteristics** | **Round 1** (*n* = 10) | **Round 2** (*n* = 7) | **Consensus meeting** (*n* = 3) |
| **Age category (years), n (%)** | | | |
| 26 - 35 | 1 (10) | 1 (14) | 1 (33) |
| 36 - 45 | 1 (10) | 1 (14) | - |
| 46 - 55 | 3 (30) | 1 (14) | 1 (33) |
| 56 - 65 | 3 (30) | 2 (29) | 1 (33) |
| 66 - 75 | 2 (20) | 2 (29) | - |
| **Gender, n (%)** | | | |
| Male | 6 (60) | 4 (57) | 3 (100) |
| Female | 4 (40) | 3 (43) | - |
| **Ethnicity, n (%)** | | | |
| Asian (Chinese) | 1 (10) | - | - |
| Asian (Japanese) | 2 (20) | 2 (29) | 2 (67) |
| White (American) | 1 (10) | - | - |
| White (Australian) | 1 (10) | 1 (14) | - |
| White (British) | 1 (10) | 1 (14) | - |
| White (Italian/Dutch) | 1 (10) | 1 (14) | 1 (33) |
| White (Swedish) | 1 (10) | - | - |
| White (Not specified) | 2 (20) | 2 (29) | - |
| **Country of residence, n (%)** | | | |
| Australia | 1 (10) | 1 (14) | - |
| England | 2 (20) | 1 (14) | - |
| Italy | 1 (10) | 1 (14) | - |
| Japan | 2 (20) | 2 (29) | 2 (67) |
| Sweden | 1 (10) | - | - |
| USA | 3 (30) | 2 (29) | 1 (33) |
| **Current academic/research role, n (%)** | | | |

| | | | |
|---|---|---|---|
| Associate professor | 3 (30) | 1 (14) | - |
| Lecturer | 2 (20) | 2 (29) | 2 (67) |
| Professor | 4 (40) | 3 (43) | 1 (33) |
| Not specified | 1 (10) | 1 (14) | - |
| **Highest educational qualification, n (%)** | | | |
| Postgraduate | 10 (100) | 7 (100) | 3 (100) |
| **Role location, n (%)** | | | |
| Hospital | 3 (30) | 2 (29) | 1 (33) |
| University | 7 (70) | 5 (71) | 2 (67) |
| **Length of academic/research experience (years), n (%)** | | | |
| 1-5 | 1 (10) | 1 (14) | - |
| 6-10 | 1 (10) | 1 (14) | 1 (33) |
| 11-15 | 2 (20) | 2 (29) | 1 (33) |
| 16 - 20 | 1 (10) | - | - |
| > 20 | 5 (50) | 3 (43) | 1 (33) |

USA: United States of America

***Not all percentages add up to 100% owing to rounding**
