## Supplementary material for "A Core Outcome Set to evaluate the impact of prognostication in people living with advanced cancer: an international consensus study": S2 File

### Rating of outcomes by stakeholder group in round 1 of the Delphi survey

| Outcomes | Patients (*n* = 10) | | | | Informal caregivers (*n* = 11) | | | | Clinicians (*n* = 18) | | | | Academics/researchers (*n* = 10) | | | | Overall (*n* = 49) | | | | Consensus |
|---|---|---|---|---|---|---|---|---|---|---|---|---|---|---|---|---|---|---|---|---|---|
| | 1-3 | 4-6 | 7-9 | UN | 1-3 | 4-6 | 7-9 | UN | 1-3 | 4-6 | 7-9 | UN | 1-3 | 4-6 | 7-9 | UN | 1-3 | 4-6 | 7-9 | UN | |
| | % | % | % | % | % | % | % | % | % | % | % | % | % | % | % | % | % | % | % | % | |
| Length of survival | 0 | 10 | 80 | 10 | 18 | 9 | 64 | 9 | 0 | 17 | 83 | 0 | 10 | 30 | 60 | 0 | 6 | 16 | 73 | 4 | |
| Pain | 0 | 10 | 90 | 0 | 0 | 18 | 73 | 9 | 6 | 28 | 61 | 6 | 10 | 50 | 40 | 0 | 4 | 27 | 65 | 4 | |
| Drowsiness | 0 | 40 | 60 | 0 | 9 | 27 | 64 | 0 | 6 | 50 | 39 | 6 | 10 | 40 | 50 | 0 | 6 | 41 | 51 | 2 | |
| Nausea | 0 | 20 | 70 | 0 | 9 | 27 | 64 | 0 | 11 | 50 | 22 | 17 | 10 | 40 | 50 | 0 | 8 | 39 | 47 | 6 | |
| General malaise | 0 | 40 | 50 | 0 | 9 | 36 | 55 | 0 | 0 | 44 | 50 | 6 | 10 | 40 | 50 | 0 | 4 | 43 | 51 | 2 | |
| Weakness | 0 | 50 | 50 | 0 | 9 | 27 | 64 | 0 | 0 | 44 | 50 | 6 | 10 | 40 | 50 | 0 | 4 | 41 | 53 | 2 | |
| Breathlessness | 0 | 40 | 60 | 0 | 9 | 36 | 55 | 0 | 0 | 44 | 44 | 11 | 10 | 10 | 70 | 10 | 4 | 35 | 55 | 6 | |
| Depression | 10 | 20 | 70 | 0 | 9 | 18 | 73 | 0 | 6 | 39 | 56 | 0 | 10 | 20 | 70 | 0 | 8 | 27 | 65 | 0 | |
| Anxiety | 0 | 20 | 70 | 0 | 0 | 9 | 82 | 9 | 6 | 33 | 61 | 0 | 10 | 20 | 70 | 0 | 4 | 24 | 69 | 2 | |
| Psychological/mental status | 10 | 30 | 50 | 0 | 9 | 9 | 82 | 0 | 6 | 22 | 61 | 11 | 0 | 30 | 70 | 0 | 6 | 24 | 65 | 4 | |
| Psychological distress | 0 | 30 | 70 | 0 | 9 | 18 | 73 | 0 | 6 | 22 | 72 | 0 | 0 | 20 | 80 | 0 | 4 | 22 | 73 | 0 | |
| Spectrum of hope | 0 | 20 | 70 | 0 | 9 | 9 | 82 | 0 | 0 | 22 | 78 | 0 | 10 | 40 | 40 | 10 | 6 | 22 | 69 | 2 | |
| Being at peace with dying | 0 | 30 | 70 | 0 | 9 | 18 | 73 | 0 | 0 | 28 | 72 | 0 | 10 | 40 | 40 | 10 | 4 | 29 | 65 | 2 | |
| Spiritual and religious coping | 10 | 30 | 40 | 10 | 18 | 9 | 73 | 0 | 0 | 44 | 50 | 6 | 20 | 50 | 30 | 0 | 12 | 35 | 49 | 4 | |
| Spiritual crisis | 20 | 50 | 20 | 0 | 18 | 27 | 55 | 0 | 6 | 33 | 56 | 6 | 0 | 50 | 50 | 0 | 12 | 39 | 47 | 2 | |
| Loss of interest/pleasure | 0 | 30 | 70 | 0 | 18 | 36 | 45 | 0 | 0 | 33 | 61 | 6 | 0 | 30 | 70 | 0 | 4 | 33 | 61 | 2 | |
| Loss of resilience | 0 | 40 | 60 | 0 | 9 | 27 | 64 | 0 | 0 | 33 | 61 | 6 | 10 | 40 | 50 | 0 | 4 | 35 | 59 | 2 | |
| Loss of dignity | 0 | 20 | 80 | 0 | 9 | 27 | 64 | 0 | 0 | 33 | 67 | 0 | 10 | 20 | 50 | 20 | 4 | 27 | 65 | 4 | |
| Dissatisfaction with life | 10 | 50 | 30 | 0 | 18 | 27 | 55 | 0 | 0 | 28 | 67 | 6 | 10 | 30 | 50 | 10 | 10 | 33 | 53 | 4 | |
| Perceived sense of burden on others | 0 | 10 | 90 | 0 | 9 | 27 | 64 | 0 | 0 | 28 | 72 | 0 | 10 | 20 | 60 | 10 | 4 | 22 | 71 | 2 | |
| Sense of suffering | 0 | 30 | 60 | 10 | 9 | 18 | 73 | 0 | 0 | 28 | 67 | 6 | 0 | 20 | 80 | 0 | 2 | 24 | 69 | 4 | |
| Sense of control | 0 | 30 | 70 | 0 | 18 | 0 | 82 | 0 | 0 | 33 | 67 | 0 | 10 | 40 | 50 | 0 | 6 | 27 | 67 | 0 | |
| Desire for death | 10 | 20 | 70 | 0 | 27 | 0 | 64 | 9 | 0 | 28 | 72 | 0 | 0 | 50 | 50 | 0 | 8 | 24 | 65 | 2 | |

| | | | | | | | | | | | | | | | | | | | | |
|---|---|---|---|---|---|---|---|---|---|---|---|---|---|---|---|---|---|---|---|---|
| Wish to live | 0 | 10 | 80 | 0 | 9 | 0 | 82 | 9 | 6 | 39 | 56 | 0 | 10 | 50 | 40 | 0 | 8 | 27 | 63 | 2 |
| Worry about dying | 10 | 10 | 70 | 0 | 18 | 0 | 82 | 0 | 0 | 28 | 72 | 0 | 0 | 20 | 80 | 0 | 6 | 18 | 76 | 0 |
| Disbelief, shock, and denial | 0 | 0 | 80 | 10 | 9 | 27 | 55 | 9 | 0 | 44 | 56 | 0 | 20 | 20 | 50 | 10 | 8 | 27 | 59 | 6 |
| Avoidance of prognosis | 10 | 10 | 70 | 0 | 18 | 36 | 36 | 9 | 6 | 44 | 50 | 0 | 20 | 30 | 50 | 0 | 14 | 33 | 51 | 2 |
| Prognostic acceptance | 10 | 10 | 70 | 0 | 18 | 27 | 55 | 0 | 0 | 39 | 56 | 6 | 10 | 20 | 70 | 0 | 10 | 27 | 61 | 2 |
| Emotional distress | 0 | 20 | 70 | 0 | 18 | 18 | 64 | 0 | 6 | 28 | 61 | 6 | 10 | 10 | 80 | 0 | 10 | 20 | 67 | 2 |
| Use of coping strategies/mechanisms | 0 | 10 | 80 | 0 | 27 | 9 | 55 | 9 | 0 | 33 | 67 | 0 | 10 | 20 | 70 | 0 | 8 | 22 | 67 | 2 |
| Fixation on prognosis | 0 | 30 | 60 | 0 | 27 | 27 | 45 | 0 | 17 | 33 | 50 | 0 | 10 | 40 | 50 | 0 | 16 | 33 | 51 | 0 |
| Mental/emotional preparation for end-of-life | 0 | 10 | 80 | 0 | 18 | 9 | 64 | 9 | 0 | 33 | 67 | 0 | 10 | 10 | 80 | 0 | 8 | 18 | 71 | 2 |
| Achieving/prioritising personal goals and values | 0 | 40 | 40 | 10 | 18 | 18 | 64 | 0 | 0 | 28 | 72 | 0 | 0 | 20 | 80 | 0 | 6 | 27 | 65 | 2 |
| Anticipatory grief in patients | 0 | 20 | 70 | 0 | 27 | 18 | 55 | 0 | 0 | 28 | 72 | 0 | 0 | 50 | 50 | 0 | 8 | 29 | 63 | 0 |
| Anticipatory grief in informal caregivers | 0 | 20 | 80 | 0 | 18 | 27 | 55 | 0 | 0 | 33 | 61 | 6 | 10 | 30 | 60 | 0 | 6 | 29 | 63 | 2 |
| Having the opportunity to say goodbye to loved ones | 10 | 10 | 80 | 0 | 27 | 0 | 73 | 0 | 0 | 33 | 67 | 0 | 10 | 10 | 80 | 0 | 10 | 16 | 73 | 0 |
| Decisional satisfaction | 0 | 0 | 90 | 0 | 18 | 9 | 64 | 9 | 0 | 50 | 50 | 0 | 10 | 20 | 70 | 0 | 6 | 27 | 65 | 2 |
| Regret in informal caregivers | 0 | 40 | 50 | 0 | 18 | 18 | 55 | 9 | 0 | 67 | 33 | 0 | 10 | 40 | 50 | 0 | 6 | 47 | 45 | 2 |
| Bereavement in informal caregivers | 0 | 20 | 60 | 10 | 9 | 18 | 73 | 0 | 6 | 50 | 44 | 0 | 0 | 30 | 70 | 0 | 4 | 35 | 59 | 2 |
| Cognitive function | 0 | 10 | 80 | 0 | 9 | 9 | 82 | 0 | 11 | 28 | 61 | 0 | 10 | 40 | 50 | 0 | 8 | 24 | 67 | 0 |
| Quality of communication between patient and family/friends | 0 | 20 | 80 | 0 | 9 | 9 | 82 | 0 | 11 | 33 | 50 | 6 | 0 | 30 | 70 | 0 | 6 | 24 | 67 | 2 |
| Quality of patient-informal caregiver relationship | 0 | 0 | 100 | 0 | 9 | 9 | 82 | 0 | 6 | 44 | 50 | 0 | 0 | 30 | 70 | 0 | 4 | 24 | 71 | 0 |
| Quality of relationships with others | 0 | 30 | 70 | 0 | 9 | 9 | 82 | 0 | 11 | 50 | 39 | 0 | 10 | 50 | 40 | 0 | 8 | 37 | 55 | 0 |
| Social isolation | 0 | 40 | 60 | 0 | 18 | 9 | 73 | 0 | 11 | 33 | 56 | 0 | 0 | 50 | 50 | 0 | 8 | 33 | 59 | 0 |
| Quality of life | 0 | 10 | 90 | 0 | 9 | 18 | 73 | 0 | 0 | 22 | 78 | 0 | 10 | 20 | 70 | 0 | 4 | 18 | 78 | 0 |
| Treatment/care preferences | 0 | 0 | 90 | 10 | 9 | 9 | 82 | 0 | 0 | 6 | 94 | 0 | 10 | 30 | 60 | 0 | 4 | 10 | 84 | 2 |
| Shared decision making | 0 | 10 | 90 | 0 | 9 | 9 | 82 | 0 | 0 | 17 | 83 | 0 | 0 | 10 | 90 | 0 | 2 | 12 | 86 | 0 |
| End-of-life/advance care planning | 0 | 20 | 80 | 0 | 9 | 9 | 82 | 0 | 0 | 17 | 78 | 6 | 20 | 10 | 70 | 0 | 6 | 14 | 78 | 2 |
| Information needs/preferences | 0 | 0 | 100 | 0 | 9 | 9 | 82 | 0 | 0 | 22 | 78 | 0 | 0 | 10 | 90 | 0 | 2 | 12 | 86 | 0 |
| Patient-doctor relationship | 0 | 10 | 90 | 0 | 9 | 0 | 91 | 0 | 0 | 33 | 61 | 6 | 0 | 20 | 80 | 0 | 2 | 18 | 78 | 2 |

| | | | | | | | | | | | | | | | | | | | | |
|---|---|---|---|---|---|---|---|---|---|---|---|---|---|---|---|---|---|---|---|---|
| Family informed about imminent death | 10 | 20 | 70 | 0 | 9 | 9 | 82 | 0 | 0 | 22 | 78 | 0 | 10 | 40 | 40 | 10 | 6 | 22 | 69 | 2 |
| Family present at time of death | 10 | 40 | 50 | 0 | 9 | 0 | 91 | 0 | 6 | 44 | 50 | 0 | 0 | 50 | 50 | 0 | 6 | 35 | 59 | 0 |
| Place of care | 0 | 10 | 90 | 0 | 9 | 0 | 91 | 0 | 6 | 33 | 61 | 0 | 10 | 30 | 60 | 0 | 6 | 20 | 73 | 0 |
| Place of death | 10 | 10 | 80 | 0 | 9 | 0 | 91 | 0 | 6 | 39 | 56 | 0 | 10 | 30 | 60 | 0 | 8 | 22 | 69 | 0 |
| Quality of death | 10 | 10 | 80 | 0 | 9 | 0 | 91 | 0 | 0 | 28 | 72 | 0 | 10 | 10 | 70 | 10 | 6 | 14 | 78 | 2 |
| Access to practical support | 0 | 0 | 100 | 0 | 18 | 0 | 82 | 0 | 0 | 33 | 67 | 0 | 0 | 60 | 40 | 0 | 4 | 24 | 71 | 0 |
| Access to financial support | 0 | 0 | 80 | 10 | 27 | 9 | 64 | 0 | 11 | 33 | 56 | 0 | 10 | 40 | 50 | 0 | 12 | 22 | 61 | 4 |
| Participation in clinical trials/research | 10 | 20 | 70 | 0 | 18 | 27 | 55 | 0 | 17 | 67 | 17 | 0 | 20 | 40 | 30 | 10 | 16 | 43 | 39 | 2 |
| Prognostic awareness | 10 | 0 | 80 | 10 | 9 | 0 | 91 | 0 | 0 | 17 | 83 | 0 | 0 | 30 | 70 | 0 | 4 | 12 | 82 | 2 |
| Prognostic understanding | 10 | 0 | 90 | 0 | 9 | 9 | 82 | 0 | 0 | 17 | 83 | 0 | 0 | 30 | 70 | 0 | 4 | 14 | 82 | 0 |
| Being aware of prognostic uncertainty | 0 | 10 | 90 | 0 | 9 | 18 | 73 | 0 | 0 | 22 | 78 | 0 | 0 | 20 | 80 | 0 | 2 | 18 | 80 | 0 |
| Practical/logistical preparation for end-of-life | 10 | 20 | 70 | 0 | 9 | 9 | 82 | 0 | 0 | 17 | 83 | 0 | 0 | 40 | 60 | 0 | 4 | 20 | 76 | 0 |
| Financial concerns | 0 | 10 | 70 | 10 | 18 | 0 | 82 | 0 | 11 | 33 | 56 | 0 | 10 | 30 | 60 | 0 | 10 | 20 | 65 | 4 |
| Hospice enrolment | 0 | 0 | 90 | 10 | 9 | 0 | 91 | 0 | 11 | 33 | 56 | 0 | 0 | 60 | 40 | 0 | 6 | 24 | 67 | 2 |
| Admission to hospital | 0 | 0 | 80 | 10 | 9 | 0 | 73 | 18 | 11 | 39 | 50 | 0 | 10 | 50 | 40 | 0 | 8 | 27 | 59 | 6 |
| Length of hospital admission | 0 | 10 | 80 | 10 | 9 | 9 | 82 | 0 | 11 | 33 | | 0 | 0 | 40 | 50 | 10 | 6 | 24 | 65 | 4 |
| Informal caregiver/family challenges | 0 | 20 | 70 | 0 | 18 | 9 | 73 | 0 | 6 | 33 | 61 | 0 | 0 | 60 | 40 | 0 | 6 | 31 | 61 | 2 |

Outcomes that reached 'consensus in' (≥70% of all participants rated the outcome as 'critical' importance (7-9) AND ≤15% as 'low' importance (1-3))

Outcomes that did not reach consensus

Outcomes that reached consensus for inclusion in each stakeholder group

UN: Unable to rate
