## Supplementary material for "A Core Outcome Set to evaluate the impact of prognostication in people living with advanced cancer: an international consensus study": S3 File

### Rating of outcomes by stakeholder group in round 2 of the Delphi survey

| Outcomes | Patients (*n* = 4) | | | | Informal caregivers (*n* = 5) | | | | Clinicians (*n* = 15) | | | | Academics/researchers (*n* = 7) | | | | Overall (*n* = 31) | | | | Consensus |
|---|---|---|---|---|---|---|---|---|---|---|---|---|---|---|---|---|---|---|---|---|---|
| | 1-3 | 4-6 | 7-9 | UN | 1-3 | 4-6 | 7-9 | UN | 1-3 | 4-6 | 7-9 | UN | 1-3 | 4-6 | 7-9 | UN | 1-3 | 4-6 | 7-9 | UN | |
| | % | % | % | % | % | % | % | % | % | % | % | % | % | % | % | % | % | % | % | % | |
| Length of survival | 0 | 0 | 100 | 0 | 0 | 20 | 80 | 0 | 0 | 13 | 87 | 0 | 0 | 14 | 86 | 0 | 0 | 13 | 87 | 0 | |
| Pain | 0 | 0 | 100 | 0 | 0 | 20 | 80 | 0 | 0 | 20 | 80 | 0 | 0 | 43 | 57 | 0 | 0 | 23 | 77 | 0 | |
| Drowsiness | 0 | 100 | 0 | 0 | 20 | 40 | 40 | 0 | 7 | 40 | 53 | 0 | 0 | 43 | 57 | 0 | 6 | 48 | 45 | 0 | |
| Nausea | 0 | 50 | 50 | 0 | 0 | 0 | 100 | 0 | 7 | 53 | 40 | 0 | 0 | 71 | 29 | 0 | 3 | 48 | 48 | 0 | |
| General malaise | 0 | 50 | 50 | 0 | 0 | 20 | 80 | 0 | 7 | 40 | 53 | 0 | 0 | 57 | 43 | 0 | 3 | 42 | 55 | 0 | |
| Weakness | 0 | 25 | 75 | 0 | 0 | 40 | 60 | 0 | 7 | 33 | 60 | 0 | 0 | 43 | 57 | 0 | 3 | 35 | 61 | 0 | |
| Breathlessness | 0 | 50 | 50 | 0 | 0 | 40 | 60 | 0 | 7 | 40 | 53 | 0 | 0 | 43 | 57 | 0 | 3 | 42 | 55 | 0 | |
| Physical functioning | 0 | 0 | 100 | 0 | 0 | 40 | 60 | 0 | 0 | 7 | 93 | 0 | 0 | 43 | 57 | 0 | 0 | 19 | 81 | 0 | |
| Depression | 0 | 25 | 75 | 0 | 0 | 40 | 60 | 0 | 0 | 20 | 80 | 0 | 0 | 43 | 57 | 0 | 0 | 29 | 71 | 0 | |
| Anxiety | 0 | 25 | 75 | 0 | 20 | 20 | 60 | 0 | 0 | 20 | 80 | 0 | 0 | 57 | 43 | 0 | 3 | 29 | 68 | 0 | |
| Psychological/mental status | 0 | 0 | 100 | 0 | 20 | 20 | 60 | 0 | 0 | 13 | 87 | 0 | 0 | 29 | 71 | 0 | 3 | 16 | 81 | 0 | |
| Psychological distress | 0 | 25 | 75 | 0 | 20 | 20 | 60 | 0 | 0 | 13 | 87 | 0 | 0 | 14 | 86 | 0 | 3 | 16 | 81 | 0 | |
| Spectrum of hope | 0 | 0 | 100 | 0 | 0 | 40 | 60 | 0 | 0 | 13 | 87 | 0 | 0 | 43 | 57 | 0 | 0 | 23 | 77 | 0 | |
| Being at peace with dying | 0 | 0 | 100 | 0 | 20 | 0 | 40 | 40 | 0 | 13 | 87 | 0 | 0 | 29 | 71 | 0 | 3 | 13 | 77 | 6 | |
| Spiritual and religious coping | 0 | 50 | 50 | 0 | 40 | 40 | 0 | 20 | 0 | 67 | 33 | 0 | 0 | 71 | 29 | 0 | 6 | 61 | 29 | 3 | |
| Spiritual/religious/existential crisis | 25 | 50 | 25 | 0 | 60 | 20 | 20 | 0 | 0 | 20 | 80 | 0 | 0 | 71 | 29 | 0 | 13 | 35 | 52 | 0 | |
| Loss of interest/pleasure | 0 | 50 | 50 | 0 | 40 | 20 | 40 | 0 | 0 | 33 | 67 | 0 | 0 | 43 | 57 | 0 | 6 | 35 | 58 | 0 | |
| Loss of resilience | 0 | 0 | 100 | 0 | 20 | 40 | 40 | 0 | 0 | 27 | 73 | 0 | 0 | 43 | 57 | 0 | 3 | 29 | 68 | 0 | |
| Loss of dignity | 0 | 0 | 100 | 0 | 20 | 0 | 80 | 0 | 0 | 7 | 93 | 0 | 0 | 29 | 71 | 0 | 3 | 10 | 87 | 0 | |
| Dissatisfaction with life | 0 | 75 | 25 | 0 | 20 | 20 | 40 | 20 | 0 | 40 | 60 | 0 | 0 | 57 | 43 | 0 | 3 | 45 | 48 | 3 | |
| Perceived sense of burden on others | 0 | 0 | 100 | 0 | 40 | 20 | 40 | 0 | 0 | 20 | 80 | 0 | 0 | 43 | 57 | 0 | 6 | 23 | 71 | 0 | |
| Sense of suffering | 0 | 0 | 100 | 0 | 20 | 20 | 60 | 0 | 0 | 13 | 87 | 0 | 0 | 29 | 71 | 0 | 3 | 16 | 81 | 0 | |
| Sense of control | 0 | 0 | 100 | 0 | 20 | 0 | 80 | 0 | 0 | 20 | 80 | 0 | 0 | 29 | 71 | 0 | 3 | 16 | 81 | 0 | |

| | | | | | | | | | | | | | | | | | | | | |
|---|---|---|---|---|---|---|---|---|---|---|---|---|---|---|---|---|---|---|---|---|
| Desire for death | 0 | 50 | 50 | 0 | 20 | 20 | 20 | 40 | 0 | 20 | 80 | 0 | 0 | 29 | 71 | 0 | 3 | 26 | 65 | 6 |
| Wish to live | 0 | 25 | 75 | 0 | 20 | 0 | 20 | 60 | 0 | 40 | 60 | 0 | 0 | 43 | 57 | 0 | 3 | 32 | 55 | 10 |
| Worry about dying | 0 | 0 | 100 | 0 | 40 | 0 | 60 | 0 | 0 | 20 | 80 | 0 | 0 | 14 | 86 | 0 | 6 | 13 | 81 | 0 |
| Disbelief, shock, and denial | 0 | 50 | 50 | 0 | 20 | 20 | 60 | 0 | 0 | 53 | 47 | 0 | 0 | 86 | 14 | 0 | 3 | 55 | 42 | 0 |
| Avoidance of prognosis | 0 | 75 | 25 | 0 | 40 | 40 | 20 | 0 | 0 | 67 | 33 | 0 | 0 | 57 | 43 | 0 | 6 | 61 | 32 | 0 |
| Prognostic acceptance | 0 | 25 | 75 | 0 | 20 | 20 | 40 | 20 | 0 | 60 | 40 | 0 | 0 | 43 | 57 | 0 | 3 | 45 | 48 | 3 |
| Emotional distress | 0 | 0 | 100 | 0 | 40 | 0 | 60 | 0 | 0 | 13 | 87 | 0 | 0 | 43 | 57 | 0 | 6 | 16 | 77 | 0 |
| Use of coping strategies/mechanisms | 0 | 0 | 100 | 0 | 40 | 0 | 60 | 0 | 0 | 33 | 67 | 0 | 0 | 43 | 57 | 0 | 6 | 26 | 68 | 0 |
| Fixation on prognosis | 0 | 75 | 25 | 0 | 40 | 40 | 0 | 20 | 0 | 53 | 47 | 0 | 0 | 71 | 29 | 0 | 6 | 58 | 32 | 3 |
| Mental/emotional preparation for end-of-life | 0 | 0 | 100 | 0 | 20 | 0 | 60 | 20 | 0 | 33 | 67 | 0 | 0 | 29 | 71 | 0 | 3 | 23 | 71 | 3 |
| Achieving/prioritising personal goals and values | 0 | 25 | 75 | 0 | 20 | 40 | 40 | 0 | 0 | 47 | 53 | 0 | 0 | 14 | 86 | 0 | 3 | 35 | 61 | 0 |
| Anticipatory grief in patients | 0 | 50 | 50 | 0 | 40 | 0 | 60 | 0 | 0 | 40 | 60 | 0 | 0 | 57 | 43 | 0 | 6 | 39 | 55 | 0 |
| Anticipatory grief in informal caregivers | 0 | 0 | 100 | 0 | 20 | 40 | 40 | 0 | 0 | 40 | 60 | 0 | 0 | 43 | 57 | 0 | 3 | 35 | 61 | 0 |
| Having the opportunity to say goodbye to loved ones | 0 | 0 | 100 | 0 | 20 | 20 | 60 | 0 | 0 | 13 | 87 | 0 | 0 | 14 | 86 | 0 | 3 | 13 | 84 | 0 |
| Decisional satisfaction | 0 | 50 | 50 | 0 | 40 | 20 | 40 | 0 | 0 | 20 | 80 | 0 | 0 | 43 | 57 | 0 | 6 | 29 | 65 | 0 |
| Regret in informal caregivers | 0 | 75 | 25 | 0 | 20 | 20 | 60 | 0 | 0 | 67 | 33 | 0 | 0 | 29 | 71 | 0 | 3 | 52 | 45 | 0 |
| Bereavement in informal caregivers | 0 | 0 | 75 | 25 | 20 | 20 | 60 | 0 | 0 | 47 | 53 | 0 | 0 | 14 | 86 | 0 | 3 | 29 | 65 | 3 |
| Cognitive function | 0 | 25 | 75 | 0 | 20 | 0 | 80 | 0 | 0 | 33 | 67 | 0 | 0 | 57 | 43 | 0 | 3 | 32 | 65 | 0 |
| Quality of communication between patient and family/friends | 0 | 0 | 100 | 0 | 20 | 0 | 80 | 0 | 0 | 27 | 73 | 0 | 0 | 14 | 86 | 0 | 3 | 16 | 81 | 0 |
| Quality of patient-informal caregiver relationship | 0 | 0 | 100 | 0 | 20 | 0 | 80 | 0 | 7 | 27 | 67 | 0 | 0 | 14 | 86 | 0 | 6 | 16 | 77 | 0 |
| Quality of relationships with others | 0 | 50 | 50 | 0 | 20 | 40 | 40 | 0 | 7 | 53 | 40 | 0 | 0 | 57 | 43 | 0 | 6 | 52 | 42 | 0 |
| Social isolation | 0 | 0 | 100 | 0 | 20 | 40 | 40 | 0 | 7 | 27 | 67 | 0 | 0 | 29 | 71 | 0 | 6 | 26 | 68 | 0 |
| Quality of life | 0 | 0 | 100 | 0 | 0 | 20 | 80 | 0 | 0 | 27 | 73 | 0 | 0 | 29 | 71 | 0 | 0 | 23 | 77 | 0 |
| Treatment/care preferences | 0 | 0 | 100 | 0 | 20 | 0 | 80 | 0 | 0 | 7 | 93 | 0 | 0 | 0 | 100 | 0 | 3 | 3 | 94 | 0 |
| Shared decision-making | 0 | 0 | 100 | 0 | 20 | 0 | 80 | 0 | 0 | 20 | 80 | 0 | 0 | 0 | 100 | 0 | 3 | 10 | 87 | 0 |
| End-of-life/advance care planning | 0 | 0 | 100 | 0 | 20 | 0 | 80 | 0 | 0 | 7 | 93 | 0 | 0 | 29 | 71 | 0 | 3 | 10 | 87 | 0 |
| Information needs/preferences | 0 | 0 | 100 | 0 | 20 | 0 | 80 | 0 | 0 | 20 | 80 | 0 | 0 | 0 | 100 | 0 | 3 | 10 | 87 | 0 |

| | | | | | | | | | | | | | | | | | | | | |
|---|---|---|---|---|---|---|---|---|---|---|---|---|---|---|---|---|---|---|---|---|
| Patient-clinician relationship | 0 | 0 | 100 | 0 | 20 | 0 | 80 | 0 | 0 | 7 | 93 | 0 | 0 | 14 | 86 | 0 | 3 | 6 | 90 | 0 |
| Family informed about imminent death | 0 | 0 | 100 | 0 | 20 | 0 | 80 | 0 | 0 | 7 | 93 | 0 | 0 | 14 | 86 | 0 | 3 | 6 | 90 | 0 |
| Family present at time of death | 0 | 25 | 75 | 0 | 20 | 0 | 80 | 0 | 7 | 27 | 67 | 0 | 0 | 14 | 86 | 0 | 6 | 19 | 74 | 0 |
| Place of care | 0 | 0 | 100 | 0 | 20 | 0 | 80 | 0 | 0 | 47 | 53 | 0 | 0 | 14 | 86 | 0 | 3 | 26 | 71 | 0 |
| Place of death | 0 | 25 | 75 | 0 | 20 | 0 | 80 | 0 | 0 | 40 | 60 | 0 | 0 | 14 | 86 | 0 | 3 | 26 | 71 | 0 |
| Quality of death | 0 | 0 | 100 | 0 | 20 | 0 | 80 | 0 | 0 | 27 | 73 | 0 | 0 | 0 | 100 | 0 | 3 | 13 | 84 | 0 |
| Access to practical support | 0 | 0 | 100 | 0 | 0 | 20 | 80 | 0 | 0 | 20 | 80 | 0 | 0 | 29 | 71 | 0 | 0 | 19 | 81 | 0 |
| Access to financial support | 0 | 25 | 75 | 0 | 20 | 20 | 60 | 0 | 0 | 47 | 53 | 0 | 0 | 43 | 57 | 0 | 3 | 39 | 58 | 0 |
| Participation in clinical trials/research | 0 | 50 | 50 | 0 | 20 | 20 | 60 | 0 | 7 | 67 | 27 | 0 | 0 | 71 | 29 | 0 | 6 | 58 | 35 | 0 |
| Prognostic awareness | 0 | 0 | 100 | 0 | 20 | 0 | 80 | 0 | 0 | 13 | 87 | 0 | 0 | 14 | 86 | 0 | 3 | 10 | 87 | 0 |
| Prognostic understanding | 0 | 0 | 100 | 0 | 20 | 0 | 80 | 0 | 0 | 13 | 87 | 0 | 0 | 14 | 86 | 0 | 3 | 10 | 87 | 0 |
| Being aware of prognostic uncertainty | 0 | 0 | 100 | 0 | 20 | 20 | 60 | 0 | 0 | 20 | 80 | 0 | 0 | 14 | 86 | 0 | 3 | 16 | 81 | 0 |
| Practical/logistical preparation for end-of-life | 0 | 0 | 100 | 0 | 20 | 0 | 80 | 0 | 0 | 0 | 100 | 0 | 0 | 29 | 71 | 0 | 3 | 6 | 90 | 0 |
| Financial concerns | 0 | 50 | 50 | 0 | 40 | 0 | 60 | 0 | 7 | 47 | 47 | 0 | 0 | 29 | 71 | 0 | 10 | 35 | 55 | 0 |
| Hospice enrolment | 0 | 75 | 25 | 0 | 20 | 20 | 60 | 0 | 0 | 40 | 60 | 0 | 0 | 29 | 71 | 0 | 3 | 39 | 58 | 0 |
| Admission to hospital | 0 | 75 | 25 | 0 | 20 | 20 | 60 | 0 | 0 | 40 | 60 | 0 | 0 | 29 | 71 | 0 | 3 | 39 | 58 | 0 |
| Length of hospital admission | 0 | 75 | 25 | 0 | 20 | 20 | 60 | 0 | 0 | 27 | 73 | 0 | 0 | 14 | 86 | 0 | 3 | 29 | 68 | 0 |
| Informal caregiver/family challenges | 0 | 0 | 100 | 0 | 20 | 20 | 60 | 0 | 0 | 53 | 47 | 0 | 0 | 43 | 57 | 0 | 3 | 39 | 58 | 0 |

Outcomes that reached ‘consensus in’ (≥70% of all participants rated the outcome as ‘critical’ importance (7-9) AND ≤15% as ‘low’ importance (1-3))

Outcomes that did not reach consensus

Outcomes that reached consensus for inclusion in each stakeholder group

UN: Unable to rate
