## Supplementary material for "A Core Outcome Set to evaluate the impact of prognostication in people living with advanced cancer: an international consensus study": S4 File

### Analysis of attrition bias in round 1 ratings in each stakeholder group

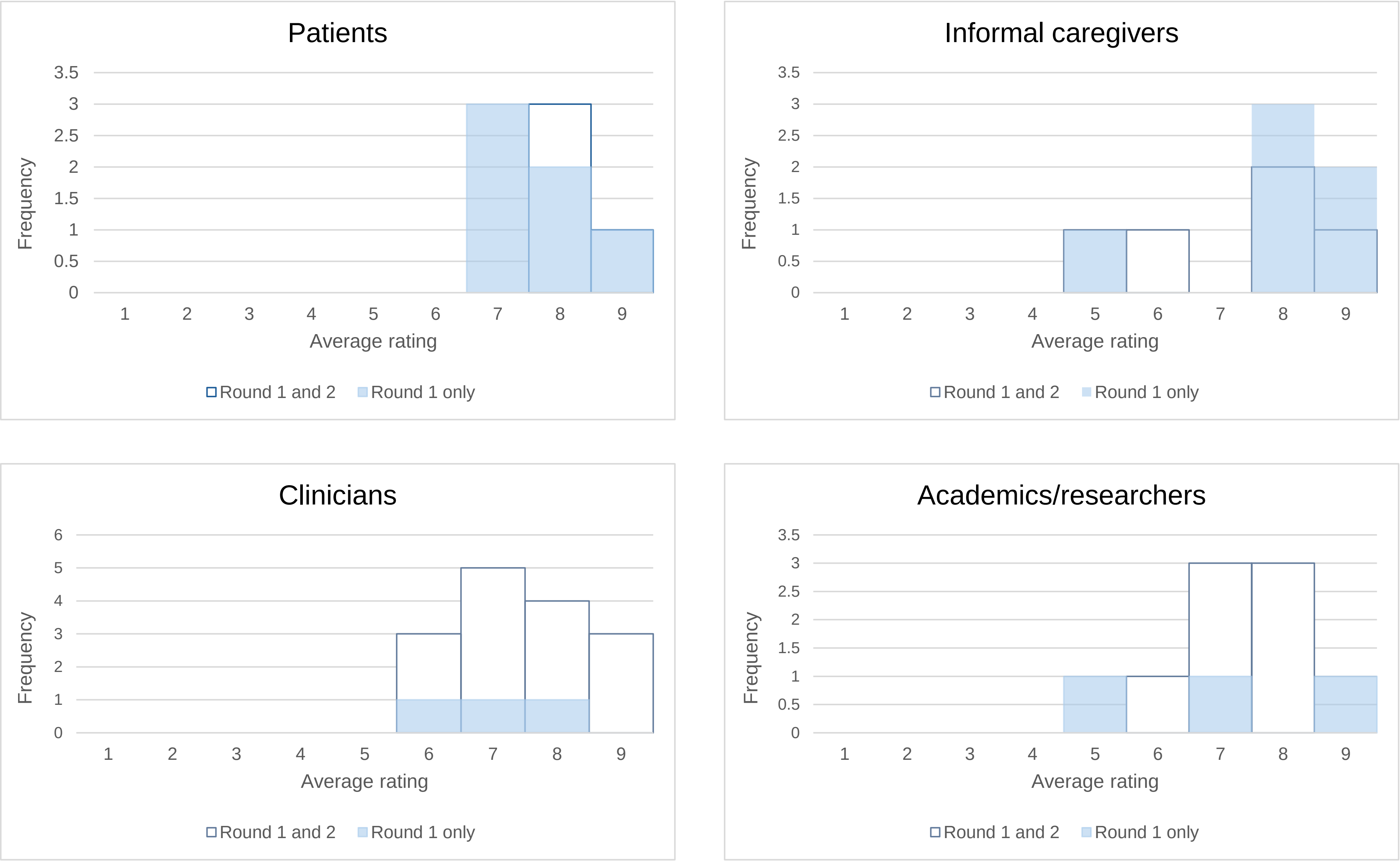

### Analysis of attrition bias in round 1 ratings for all participants

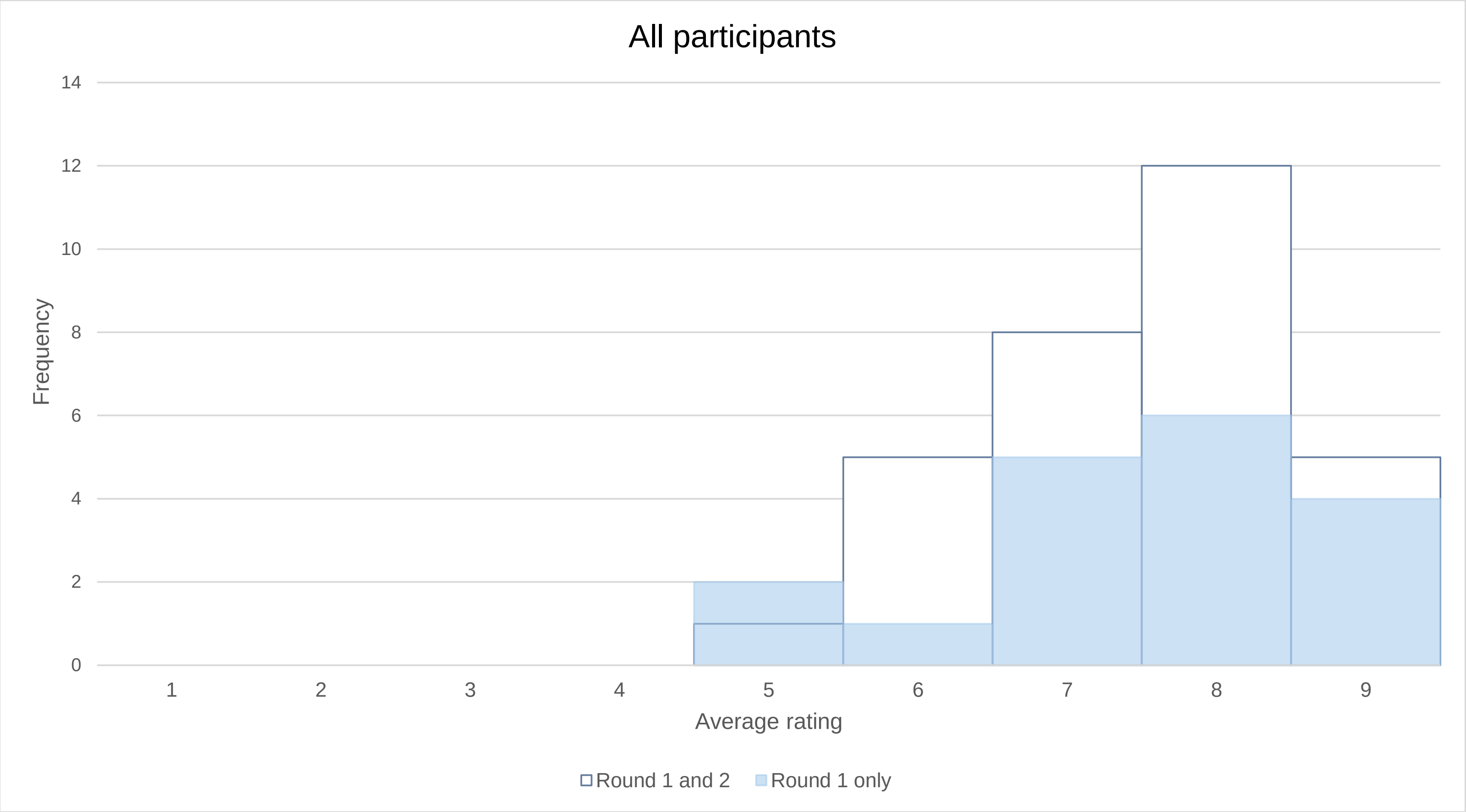
