## Supplementary material for "A Core Outcome Set to evaluate the impact of prognostication in people living with advanced cancer: an international consensus study": S5 File

### Rating of outcomes in the consensus meeting

| Outcomes | 1-3 (%) | 4-6 (%) | 7-9 (%) | Consensus |
|---|---|---|---|---|
| Length of survival | 17 | 8 | 75 | Consensus out |
| Pain | 36 | 0 | 64 | Consensus out |
| Physical functioning | 8 | 17 | 75 | Consensus in |
| Depression | 45 | 9 | 45 | Consensus out |
| Psychological/mental status | 8 | 17 | 75 | Consensus in |
| Psychological distress | 25 | 17 | 58 | Consensus out |
| Spectrum of hope | 42 | 17 | 42 | Consensus out |
| Being at peace with dying | 50 | 25 | 25 | Consensus out |
| Loss of dignity | 33 | 17 | 50 | Consensus out |
| Perceived sense of burden on others | 17 | 8 | 75 | Consensus out |
| Sense of suffering | 42 | 17 | 42 | Consensus out |
| Sense of control | 33 | 17 | 50 | Consensus out |
| Worry about dying | 33 | 8 | 58 | Consensus out |
| Emotional distress | 42 | 8 | 50 | Consensus out |
| Mental/emotional preparation for end-of-life | 17 | 0 | 83 | Consensus out |
| Having the opportunity to say goodbye to loved ones | 33 | 0 | 67 | Consensus out |
| Quality of communication between patient and family/friends | 17 | 8 | 75 | Consensus out |
| Quality of patient-informal caregiver relationship | 33 | 8 | 58 | Consensus out |
| Quality of life | 8 | 8 | 83 | Consensus in |
| Treatment/care preferences | 8 | 0 | 92 | Consensus in |

| | | | | |
|---|---|---|---|---|
| Shared decision making | 25 | 8 | 67 | Consensus out |
| End-of-life/advance care planning | 8 | 0 | 92 | Consensus in |
| Information needs/preferences | 64 | 8 | 36 | Consensus out |
| Patient-clinician relationship | 25 | 0 | 75 | Consensus out |
| Family informed about imminent death | 33 | 0 | 67 | Consensus out |
| Family present at time of death | 58 | 8 | 33 | Consensus out |
| Place of care | 8 | 0 | 92 | Consensus in |
| Place of death | 33 | 0 | 67 | Consensus out |
| Quality of death | 8 | 0 | 92 | Consensus in |
| Access to practical support | 33 | 0 | 58 | Consensus out |
| Prognostic awareness | 25 | 8 | 67 | Consensus out |
| Prognostic understanding | 8 | 0 | 92 | Consensus in |
| Being aware of prognostic uncertainty | 33 | 0 | 67 | Consensus out |
| Practical/logistical preparation for end-of-life | 8 | 8 | 83 | Consensus in |
